# Responsiveness to risk explains large variation in COVID-19 mortality across countries

**DOI:** 10.1101/2020.12.11.20247924

**Authors:** Tse Yang Lim, Hazhir Rahmandad

## Abstract

**Background:** Health outcomes from the COVID-19 pandemic vary widely across countries – by 2022 New Zealand suffered ∼10 total deaths per million people, whereas the U.K. and U.S. have had over 2000. Differences in infection fatality rates are insufficient to explain such vastly divergent outcomes. We propose that endogenous behavioral responses to risk shape countries’ epidemic trajectories, and that differences in responsiveness to risk are a primary driver of variation in epidemic scale and resultant mortality.

**Methods:** We develop several testable predictions based on the proposed endogenous risk response mechanism. We test these using a simple modified SEIR model incorporating this mechanism, which we estimate for 131 countries (5.96 billion people) using data on daily reported SARS-CoV-2 infections and COVID-19 deaths. We further examine associations between COVID-19 deaths and several observed and model-estimated country characteristics using linear regression.

**Findings:** We find empirical support for all predictions tested: 1) endogenous risk response substantially improves an SEIR model’s fit to data (mean absolute errors normalized by mean=66% across countries, vs. 551% without endogenous risk response); 2) *R*_*e*_ converges to ∼1 across countries in both empirical data and model estimates with endogenous risk response (but not without it); 3) most cross-country variation in death rates cannot be explained by intuitively important factors like hospital capacity or policy response stringency; and 4) responsiveness to risk, which governs the sensitivity of the endogenous risk response, correlates strongly with death rates (R^2^=0.75) and is the strongest explanatory factor for cross-country variation therein.

**Interpretation:** Countries converge to policy measures consistent with *R*_*e*_∼1 (or exponentially growing outbreaks will compel them to increase restrictions). Responsiveness to risk, i.e. how readily a country adopts the required measures, shapes long-term cases and deaths. With greater responsiveness, many countries could considerably improve pandemic outcomes without imposing more restrictive control policies.

**Funding:** None.

**Research in context:** *Evidence before this study:* While numerous studies have examined drivers of variation in COVID-19 infection fatality rates (e.g. age, comorbidities), relatively few have sought to explain cross-country variation in overall mortality outcomes. Mortality outcomes are shaped primarily by variation in infection rates and only secondarily by IFR variation, yet the drivers of cross-country variation in infection rates are poorly understood as well. There is ample evidence that policy responses such as mask mandates, quarantines, and other non-pharmaceutical interventions (NPIs) can influence transmission, yet across countries the association of policy responses with long-term infection rates remains weak.

*Added value of this study:* We propose a simple and intuitive theoretical mechanism – endogenous behavioral response to risk – to explain variation in cases and deaths across countries. While simple, the implications of this mechanism are rarely examined; a recent review found only 1 of 61 models in the CDC COVID-19 Forecast Hub incorporates endogenous risk response. We demonstrate how it can explain several empirical regularities, including multiple outbreak waves and convergence in effective reproduction number, which existing models largely do not explain. Our empirical estimates of responsiveness to risk also show that it drives wide cross-country variation in mortality outcomes, which otherwise remains unexplained.

*Implications of all the available evidence:* Our results highlight the key role of responsiveness to risk in shaping COVID-19 mortality outcomes. They suggest that adopting similar policy responses but with greater responsiveness could improve outcomes and reduce mortality. The responsiveness mechanism also resolves the apparent paradox that despite clear evidence of proximal NPI effectiveness, their impact on infection and death outcomes in cross-country comparisons is rather weak. Our results highlight the need to understand better the determinants of differences in responsiveness across countries, and how to improve it.

## Introduction

A single disease, COVID-19, has resulted in per-capita infection and death rates that vary by more than two orders of magnitude across nations ^1^. What explains this wide variation in outcomes?

Fundamentally, COVID-19 mortality rates are a function of two factors – infection fatality rates (IFRs) and the overall scale of the epidemic. Several factors are known to influence COVID-19 IFRs, including population age structure, prevalence of comorbidities such as cardiovascular disease or obesity, and availability of hospital or healthcare capacity ^2–8^. But across countries, the variation in IFRs resulting from these factors is far smaller than – and only weakly correlated with – the variation in overall per-capita death rates ^9,10^. Differences in ascertainment rates may explain some variation in reported disease burdens ^11^, but controlling for under-reporting of cases and deaths based on testing rates still yields wide inter-country variation ^12^. Moreover, considerable heterogeneity exists even in the more reliable death data ^12^.

Thus, to understand mortality differences, we need to understand the variation in the overall scale of the epidemic. One may expect those differences to be due to different policy measures, e.g. social distancing, mask mandates, and closures that countries have adopted to contain outbreaks ^13^. Yet the range of available policy options is similar across countries, with only a weak correlation between stringency of restrictions and national case or mortality burden ^14^. Furthermore, indices of pandemic preparedness or healthcare capacity show little relationship with COVID-19 outcome variation across countries ^8^.

We propose a simple unified mechanism to explain the wide variation in cases and deaths across countries despite broadly similar policy responses. Specifically, we argue that differences in *responsiveness to risk* across countries can lead to widely varying levels of morbidity and mortality. We first develop this argument into testable predictions, then demonstrate its value using a simplified model of COVID-19 incorporating an endogenous behavioral risk response, which closely explains observed death rates across 131 countries constituting 5.96 billion people.

### Risk responses and outbreak trajectories

In an uncontrolled epidemic, infections spread unchecked in a single massive outbreak, slowing only when most of the susceptible population has been depleted ^15,16^. With COVID-19, however, outbreaks triggered voluntary and mandatory policy and behavioral responses (non-pharmaceutical interventions: NPIs), which helped bring those outbreaks under control ^14,17–20^. As outbreaks receded, NPIs were gradually relaxed, enabling new waves^21^. The idea that those responses to risk matter is widely accepted. However, its full implications only emerge when the mechanism is modeled as an endogenous feedback process in which epidemic and societal behaviors co-evolve: behavior modifies transmission, which shapes mortality, and thus feeds back to change risk perceptions and behaviors, in turn moderating future transmission ^22,23^. With a few exceptions ^12,24,25^, this endogenous feedback mechanism is missing from current models. A recent review of models in CDC’s COVID-19 forecast hub finds only 1/61 models captures the endogenous feedback ^26^.

This endogenous mechanism shapes the trajectory of the effective reproduction number, *R*_*e*_. Outbreaks grow exponentially when *R*_*e*_ exceeds 1, increasing deaths and perceived risk, until the community adopts enough NPIs to bring down *R*_*e*_ below 1, lowering transmissions and deaths ^17,18,27,28^. Conversely, *R*_*e*_ values below 1 reduce infections and perceived risk, leading to relaxation of NPIs, which allows *R*_*e*_ back up ^29^. With responses strengthening as outbreaks grow (*R*_*e*_ > 1) and easing when they slow down (*R*_*e*_ < 1), average *R*_*e*_ in any given country converges towards one ^21^. Convergence in *R*_*e*_ may not be smooth; delays in changing risk perception and responses mean *R*_*e*_ may oscillate around 1, creating the pattern of waves seen across many countries.

### Implications for epidemic scale and mortality

Endogeneity links the strength of responses to the perceived severity of outbreaks. We approximate this linkage by relating some measure of perceived severity, e.g. perceived death rates, with some measure of response strength. *Responsiveness to risk* is then the parameter[s] that govern the strength of the relationship – a more responsive country (state, community, etc.) will adopt stronger responses at lower perceived severity, whereas a less responsive country will adopt similar measures only at higher perceived risk. For example, in late 2020 to early 2021, Australia, Hong Kong, and the UK all (re-)imposed large-scale school closures in response to renewed outbreaks. While similar in magnitude, these responses occurred when reported daily infections were at approximately 0.2, 11, and 800 new cases per million people ^1^, indicating wide variation in *responsiveness*.

Barring large differences across countries in susceptible fraction or in basic reproduction number *R*_*0*_, the convergence of *R*_*e*_ to ∼1 due to endogenous risk responses implies that different countries tend to converge to roughly similar reductions in infectious contact rates, at least prior to vaccine deployment. Countries may vary in specific policy responses, e.g. large-scale lockdowns, contact tracing & targeted quarantines, mask mandates, etc., but the strength of responses in terms of net effect on transmission rates are roughly comparable and correspond to strengths needed to bring *R*_*e*_ to ∼1. Yet countries may vary widely in their *responsiveness*, i.e. imposing the necessary (and similar) level of response at very different epidemic severities. As a result, despite convergence in terms of responses, countries may diverge on health outcomes such as mortality rates.

The proposed endogenous risk response mechanism thus yields several testable predictions (P1-4):

P1) **Improved Fit**: Incorporating an endogenous behavioral risk response will enhance the ability of an epidemic model to fit observed data.

P2) ***R***_***e***_**∼1**: Across different countries, on timescales of several months or longer, the endogenous risk response will drive convergence in *R*_*e*_ to ∼1.

P3) **Response**≠**Fewer Deaths**: On longer timescales, the average magnitude of response will be largely uncorrelated with average mortality.

P4) **Responsiveness**∼**Fewer Deaths:** Conversely, *responsiveness to risk* will vary widely across different countries, and this variation will explain much of the inter-country variation observed in long-term average mortality.

## Methods

To test these predictions, we develop a simplified epidemic model that accounts for behavioral change in response to risk. We then estimate this model using data from different countries to assess how well theoretical predictions from the model correspond to empirical regularities.

Classical epidemic models, such as the Susceptible-Infectious-Recovered (SIR) model, do not account for behavioral and policy responses to outbreaks. Contemporary models often incorporate such responses exogenously – as a function of time; using historical data on policies adopted; or as policy switches that are activated in response to risks ^28,30–32^. A few studies have instead treated responses endogenously, using behavioral ^12,24,33^ or rational ^34,35^ functions relating risks to contact rates. Our model builds upon the latter approach.

This model is simple by design, intended only to highlight the generatively sufficient role of a simple risk response mechanism in explaining large variation in outcomes. We thus exclude such factors as hospitalization, treatment capacity, testing, vaccines, variants, travel, heterogeneities in demographics and risk factors or disease severity, and many more.

### Model specification

We use a classic compartmental SEIR (susceptible-exposed-infectious-recovered) model ^15^ with one primary modification to incorporate behavioral responses that endogenously change contact rates as a continuous function of perceived risk, detailed below; see S1 for full model specification.

As in a standard SEIR model, infection rates (*r*_*E*_) depend on the susceptible (S) and infectious (I) relative to total (N) populations, and daily infectious contacts per case (*β*):

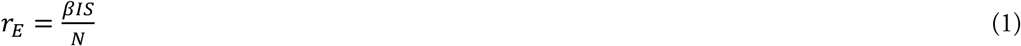

We model the effects of behavioral and policy responses to risk by allowing infectious contact rate *β* to go below its initial value *β*_0_ in response to perceived risk of death (*D*):

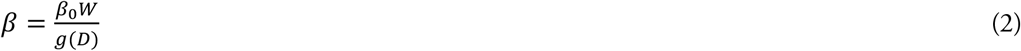

The response function *g* reflects the strength of responses affecting contact rates. *g(D)*=1 indicates pre-pandemic behavior, whereas full societal lockdown may push *g(D)* to large values, bringing *β* to ∼0. *W* is the seasonal effect of weather on COVID-19 transmission, estimated elsewhere ^12,36^. We use a simple linear form for *g*:

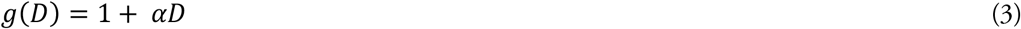

α represents the responsiveness of a community to risk; α=0 recovers the basic SEIR model. *D* is modeled as an exponential average of reported per-capita daily death rate, with an asymmetric adjustment time for increasing vs. decreasing perceived risk; see S1. To capture potential changes in risk-response relationship over time (e.g. due to adherence fatigue), we allow responsiveness α to vary linearly with time:

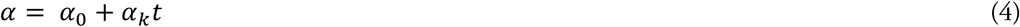

We assume a 4-day latent period and 10-day infectious period ^37,38^, calculate base IFR for each country based on age distribution ^4^ (see S3a), and use prior estimates for *W* ^12,36^. Remaining model parameters are estimated (see **Table S1**).

### Data and Estimation

We estimate the model using 7-day rolling averages of daily reported COVID-19 infections and deaths ^1^. We also use several other country statistics for analysis of results (but not model estimation; see **Table S2**). We estimate parameters separately for all 131 countries for which sufficient data are available, covering 5.96 billion people. We limit the estimation period to 31 Dec 2019 to 31 Mar 2021, exluding the effects of vaccination and new variants. While important for ongoing projections, capturing those effects adds much complexity to exposition with limited additional insight.

Estimation is by maximizing the likelihood of observing reported cases and deaths, using a negative binomial likelihood function to accommodate heteroskedasticity and fat tails (see S2). For each country we also estimate the arrival time of the first patient (*t*_*Z*_) and expected variability of the data for cases and deaths (*λ*_*i*_, *λ*_*d*_), which informs the likelihood function. To account for under-ascertainment, we specifically predict reported cases, while the model endogenously simulates the true magnitude of the epidemic in each country, by assuming the extent of under-ascertainment is relatively stable on the timescale of the disease duration (∼two weeks), even if it changes over longer time horizons (see S1a).

We also estimate the model for all countries a second time with *α* = 0, inactivating the response function and recovering the best-fit SEIR model with no risk response (henceforth NRR).

## Results

This simple model replicates well the diverse range of trajectories observed in over a year of the epidemic (**Figure 1**). Over the full sample of 131 countries, average R^2^ for infections and deaths against data is 0.56, while average of the mean absolute errors normalized by mean (MAEN) is 66%. With so simplified a model, goodness of fit is limited, but still far better than the base SEIR model without risk response (NRR R^2^=0.36, MAEN=551%) – supporting P1 (**Improved Fit**).

**Figure 1.**
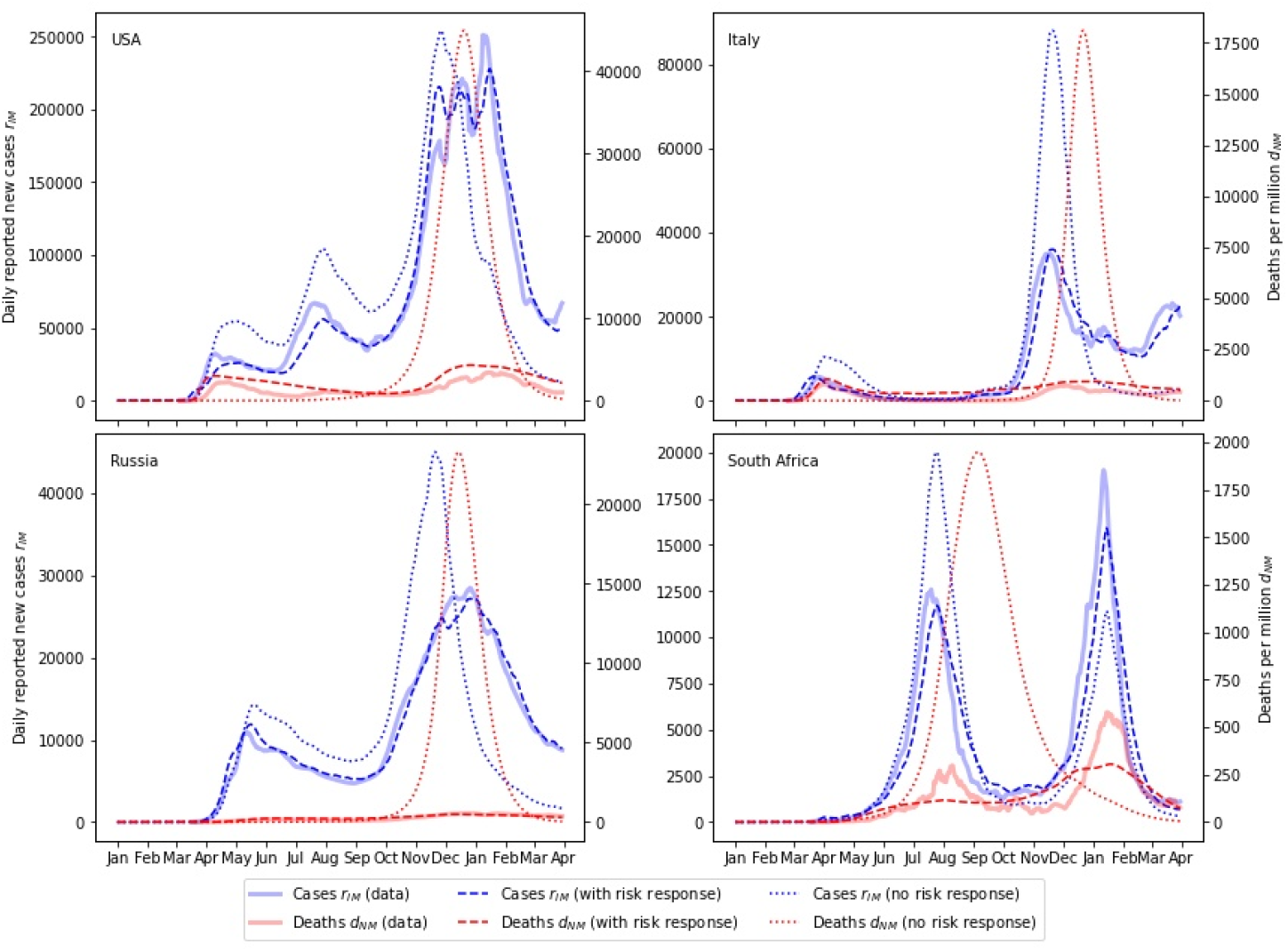
Model fit to data for selected countries, for reported cases (blue) and deaths (red). Simulated results for the model incorporating endogenous risk response (dashed) fit the data (solid) far better than the NRR model, which excludes endogenous risk response (dotted). In particular, the main model replicates multiple waves of deaths, while the NRR model cannot.

Consistent with P2 (***R***_***e***_**∼1**), over time horizons of weeks to months, estimated *R*_*e*_ across all 131 countries converges to approximately 1 (**Figure 2**; median *R*_*e*_ over 6 months prior to 31 Mar 2021 = 1.02, 90% range 0.95-1.21). Our estimated *R*_*e*_ values are comparable to independent estimates ^39^ (**Figure 2**), which similarly converge to ∼1 (median *R*_*e*_=1.05, 90% 0.93-1.14). Notably, however, NRR model estimates of *R*_*e*_ do not show similar convergence (median *R*_*e*_=0.88, 90% 0.42-1.25).

**Figure 2.**
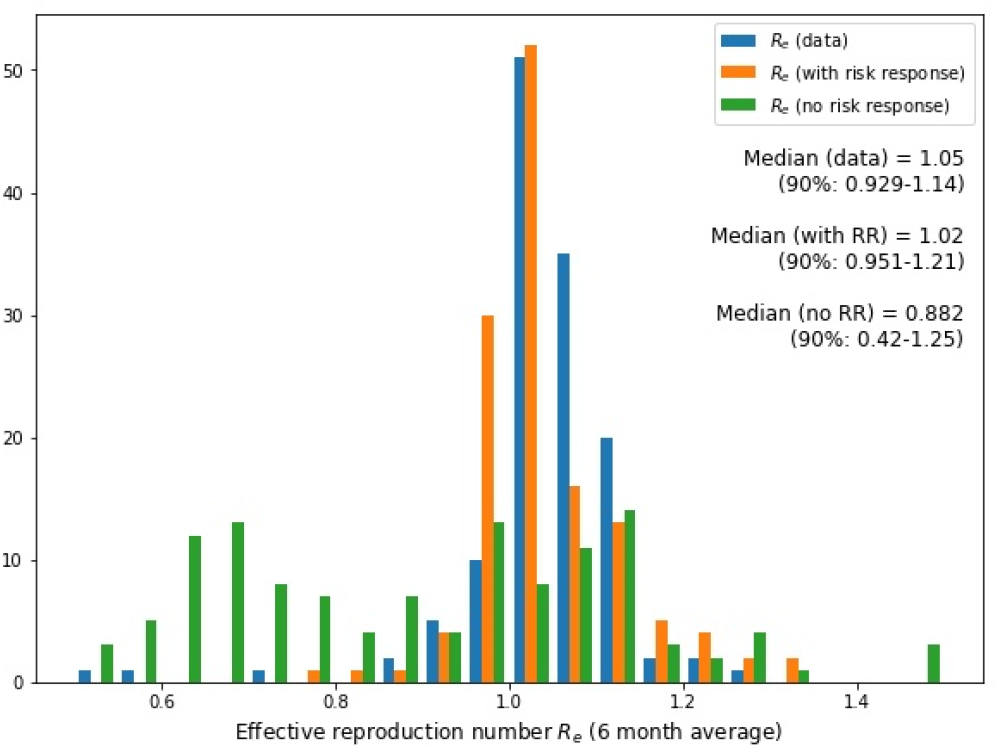
Convergence in effective reproduction number R_e_, averaged over the last six months to 31 Mar 2021. Both simulated (orange) and independently estimated (blue) R_e_ values converge to ∼1, but not in the NRR model (green).

While *R*_*e*_ has largely converged across countries to ∼1, average per-capita death rates continue to vary across countries by 2-3 orders of magnitude (90% range: 0.02-8.74 deaths per million people per day) (**Figure 3**). To test P3, we correlate variation across countries in per capita death rates averaged over the six months ending at 31 Mar 2021 with several factors that, intuitively, may influence mortality outcomes (**Figure 4**).

**Figure 3.**
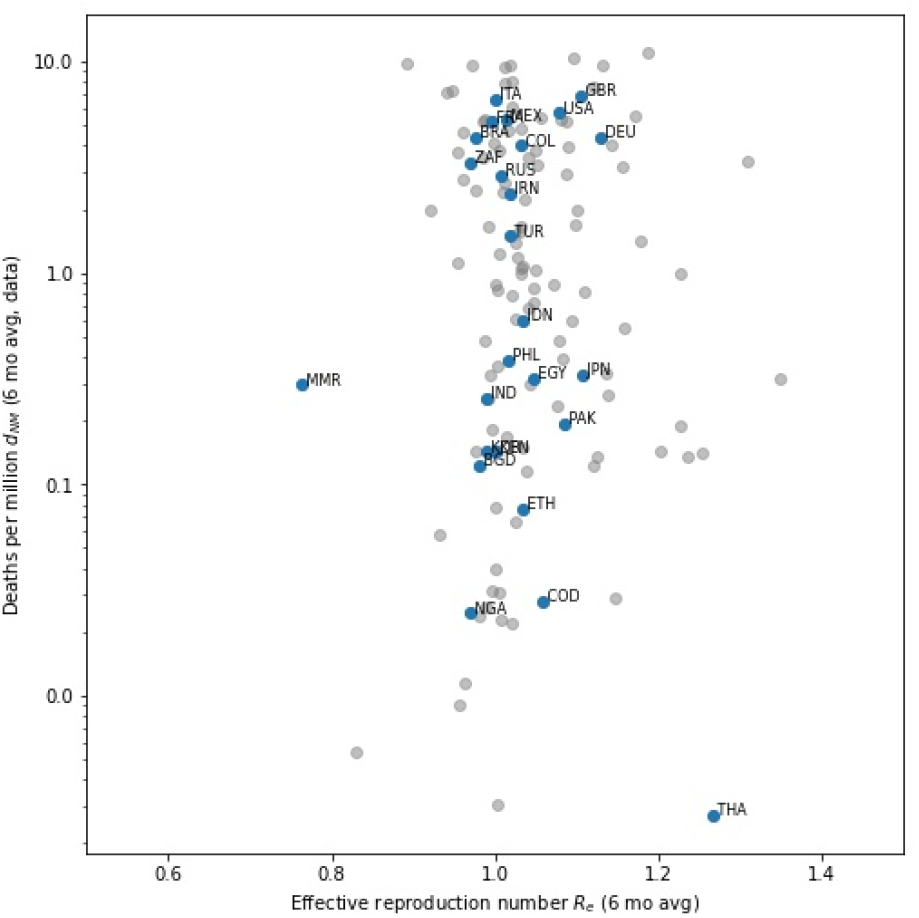
Reported death rates per million people against effective reproduction number, both averaged over the last six months to 31 Mar 2021. While R_e_ has largely converged to ∼1, death rates continue to vary by 2-3 orders of magnitude (90%: 0.02-8.74 deaths per million people per day).

**Figure 4.**
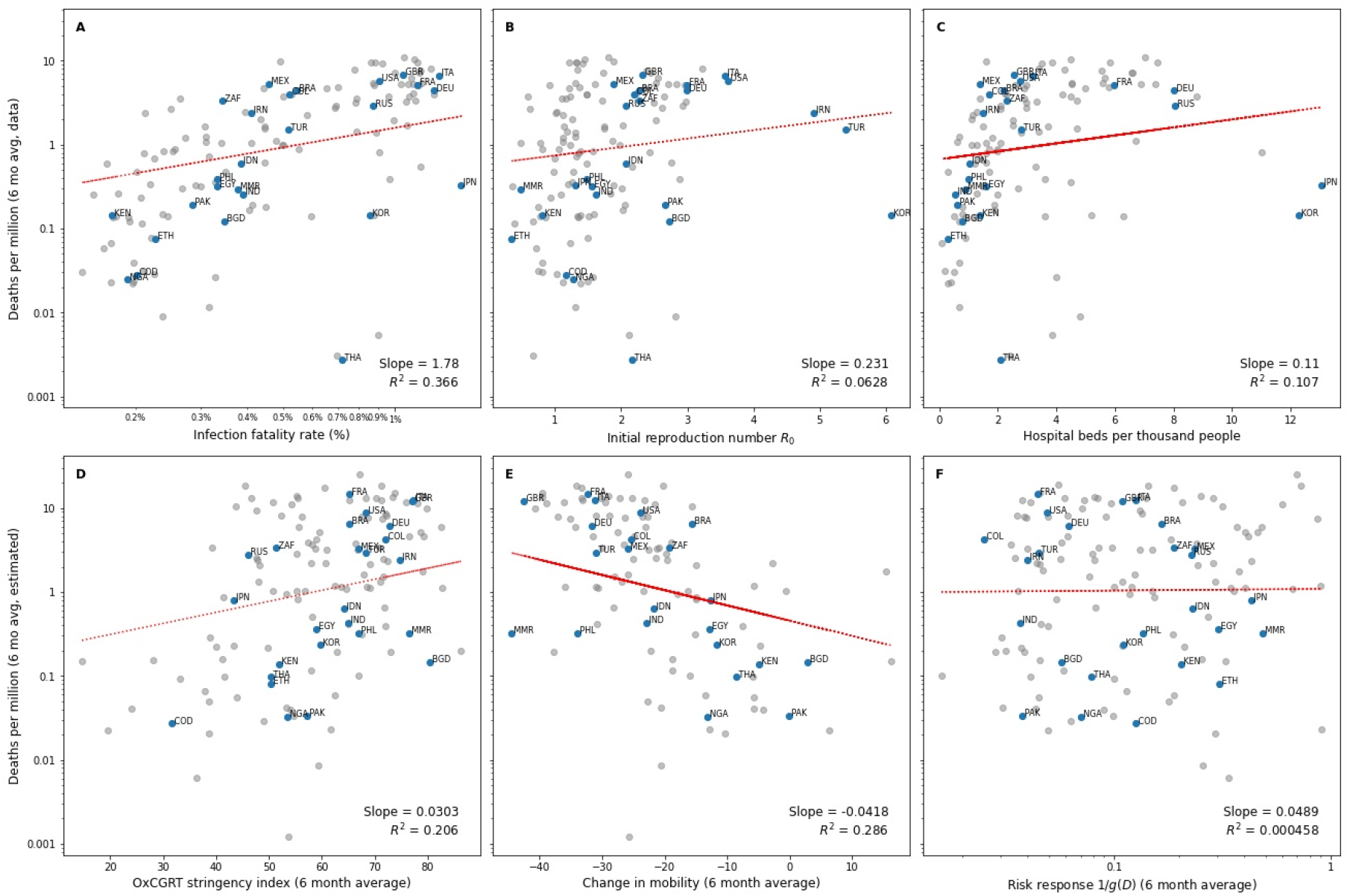
Reported death rates per million people correlated with alternative explanatory factors: A) age-adjusted infection fatality rate (note the log scale for IFR); B) initial reproduction number R_0_; C) hospital beds per thousand people; D) response stringency data; E) change in mobility relative to pre-pandemic levels; and F) estimated risk-driven change in contacts (1/g(D)).

The strongest correlation is with countries’ age-adjusted IFRs (R^2^=0.37, **Figure 4**A); unsurprisingly, countries with higher expected IFRs also have higher death rates. However, IFR varies across countries by only about 10x (90% range: 0.17%-1.2%), far less than the 2-3 orders of magnitude variation in overall per-capita death rates. Substantial variation in death rates should therefore be understood as primarily due to the scale of cases, not IFR differences.

Death rates are largely uncorrelated with initial reproduction numbers *R*_*0*_ (**Figure 4**B). Initial *R*_*0*_ reflects a diverse range of social, cultural, and geographic factors influencing transmission rates in a country’s ‘default’ state. Note that the values shown are the first available values for each country from independent estimates of *R* ^39^. Besides ‘true’ variability in *R*_*0*_, these may incorporate early behavioral or policy responses to the threat of COVID-19, and are thus lower than true basic reproduction number estimates ^40^. Nevertheless, initial variations in *R*_*0*_ are modest (90% range: 0.65-2.99) and weakly correlated with deaths.

Death rates are positively correlated with hospital beds per thousand people (a proxy for healthcare capacity; **Figure 4**C), offering no support for healthcare capacity as a prime explanation. **Figure S3** shows similarly limited correlations with other potential explanatory factors: per capita GDP, median age, and visits to retail & recreation and workplaces.

Direct tests of P3 (**Response**≠**Fewer Deaths**) compare how societal responses correlate with death rates. We consider three such measures. First, the stringency of governmental policy response aimed at reducing infectious contact rates (per the Oxford Covid-19 Government Response Tracker stringency index ^13^), e.g. school closures, travel restrictions, etc., is *positively* correlation with per capita death rates (**Figure 4**D). Second, greater reductions in mobility from pre-pandemic levels likewise correlate with *higher* death rates (**Figure 4**E, **Figure S4**). While more stringent policies and greater mobility reductions might be expected to reduce outbreak severity ^17,28,32^, endogeneity of risk responses means more severe outbreaks trigger stronger policies ^14^ and greater reductions in mobility, which could reverse the expected correlation. Regardless of endogeneity, neither measure offers a plausible explanation for variation in deaths, supporting P3. Third, while countries varied somewhat in the estimated extent of contact reduction due to their risk responses (1/*g(D)*), those reductions are uncorrelated with death rates (**Figure 4**F), further reinforcing P3.

Finally, we test P4 (**Responsiveness∼Fewer Deaths**) by correlating responsiveness and death rates. Estimated responsiveness varies widely across countries. Increasing daily deaths per million from 0 to 1 would reduce infectious contacts by 18% in the median country, with substantial between-country variation (90% range: 0.2-83% contact reduction). Responsiveness is strongly negatively correlated with 6-month averaged death rates (R^2^=0.75, **Figure 5**), supporting P4.

**Figure 5.**
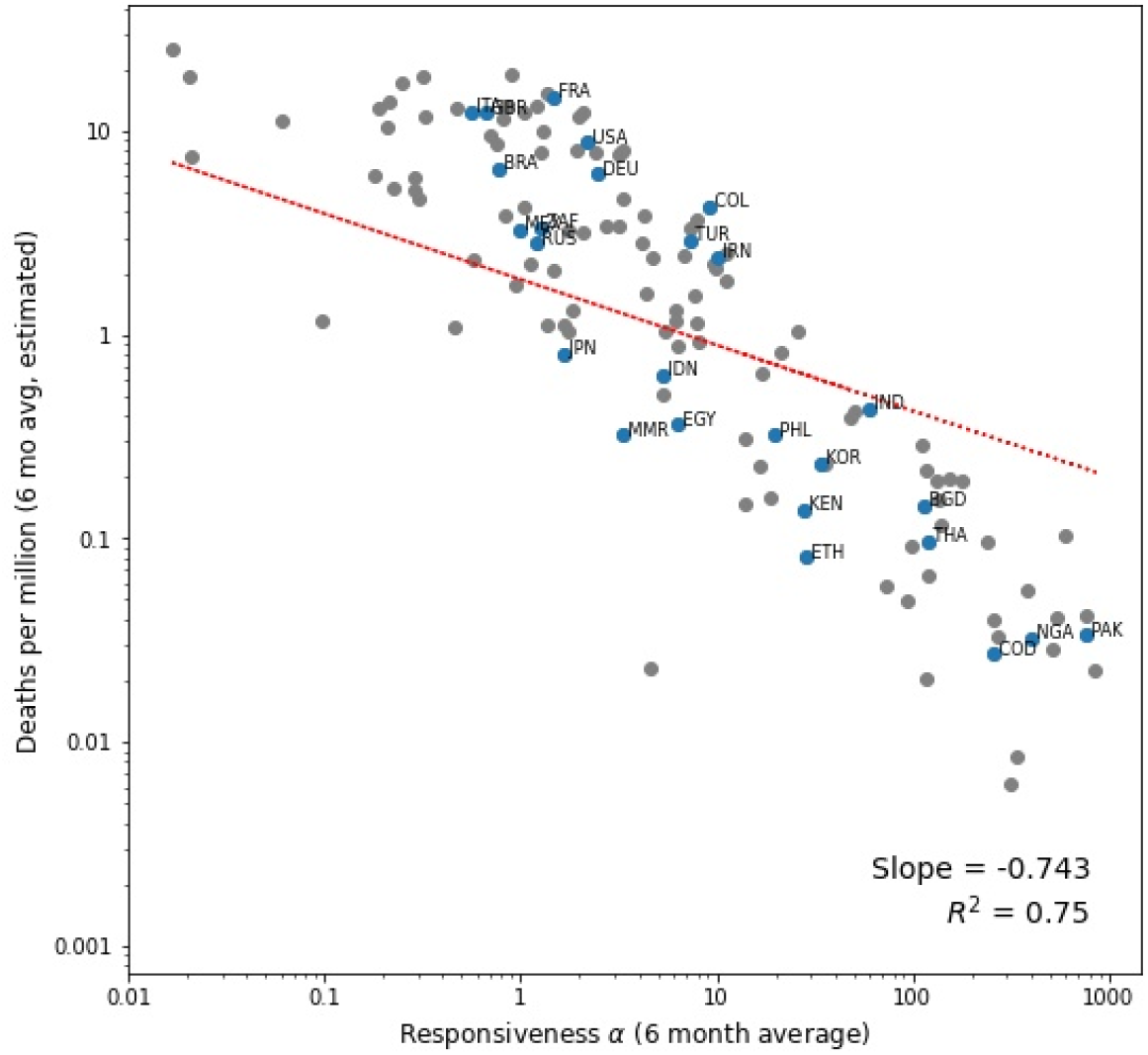
Estimated death rates per million people against responsiveness α, both averaged over six months preceding 31 Mar 2021, showing a clear negative correlation.

As a further test of P3 and P4, we regress 6-month averaged death rates against several predictive factors simultaneously (**Table 1**) to account for correlations among explanatory factors. Consistent with P4, responsiveness (log10 α) is the most important driver of variation in death rates (*t*=-11.3, *p*=5.6E-20), enhancing the model’s fit (adj. R^2^) by 0.284 (from 0.47 to 0.76); increasing responsiveness by one standard deviation reduces death rates by four times (0.24 (0.17-0.34)). In comparison, while both (log10) IFR and (log10) GDP per capita are significant predictors of death rates (*t*=2.7, *p*=0.008 and *t*=2.3, *p*=0.022 respectively), their contributions to model fit (adj. R^2^) are substantially smaller (0.014, 0.010); increasing IFR or GDP by one standard deviation increases death rates by 1.25 (1.03-1.52) times or 1.26 (0.98-1.61) times respectively. Other factors such as hospital capacity and response stringency have no significant effect and do not explain variation in death rates, further supporting P3.

**Table 1.**
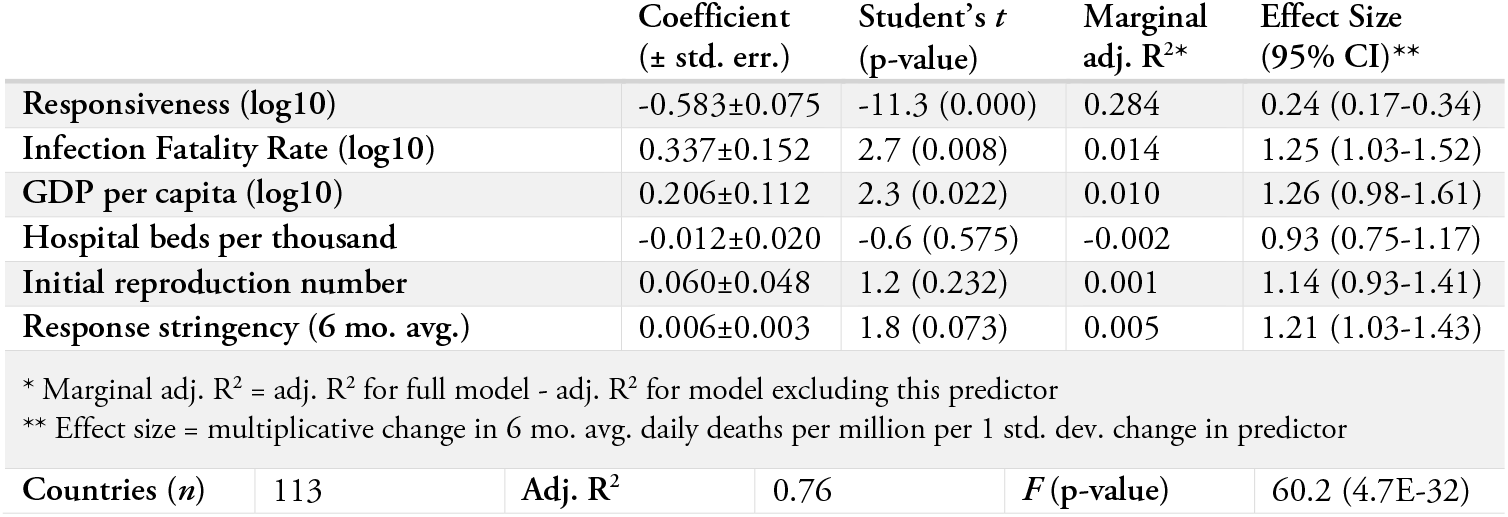
Predictors of cross-country variation in mortality rates per capita.

| | | Coefficient<br>(± std. err.) | Student's $t$<br>(p-value) | Marginal<br>adj. $R^2$* | Effect Size<br>(95% CI)** |
| --- | --- | --- | --- | --- | --- |
| **Responsiveness (log10)** | | -0.583±0.075 | -11.3 (0.000) | 0.284 | 0.24 (0.17-0.34) |
| **Infection Fatality Rate (log10)** | | 0.337±0.152 | 2.7 (0.008) | 0.014 | 1.25 (1.03-1.52) |
| **GDP per capita (log10)** | | 0.206±0.112 | 2.3 (0.022) | 0.010 | 1.26 (0.98-1.61) |
| **Hospital beds per thousand** | | -0.012±0.020 | -0.6 (0.575) | -0.002 | 0.93 (0.75-1.17) |
| **Initial reproduction number** | | 0.060±0.048 | 1.2 (0.232) | 0.001 | 1.14 (0.93-1.41) |
| **Response stringency (6 mo. avg.)** | | 0.006±0.003 | 1.8 (0.073) | 0.005 | 1.21 (1.03-1.43) |
| * Marginal adj. $R^2$ = adj. $R^2$ for full model - adj. $R^2$ for model excluding this predictor | | | | | |
| ** Effect size = multiplicative change in 6 mo. avg. daily deaths per million per 1 std. dev. change in predictor |  |  |  |  |  |
| **Countries ($n$)** | 113 | **Adj. $R^2$** | 0.76 | **$F$ (p-value)** | 60.2 (4.7E-32) |

## Discussion

The same disease, COVID-19, has resulted in mortality burdens that vary by over 100x across nations. We proposed that understanding this heterogeneity requires an endogenous view, where societal responses to the pandemic co-evolve with its severity and responsiveness to risk determines what levels of perceived severity are required to elicit effective responses. This view explains several important regularities. First, as the pandemic unfolded, all communities adopted various NPIs to curb its spread as the severity of outbreaks grew. These NPIs brought down effective reproduction numbers, lowering transmission rates. Then, as outbreaks receded, relaxation of these responses allowed new waves to emerge. The endogenous adjustment of responses to changing risks can explain the pattern of multiple outbreak waves observed across nations, and significantly enhances the ability of a simple SEIR model to fit observed patterns of infections and deaths (Prediction P1). Furthermore, this mechanism results in a longer-term convergence in *R*_*e*_ to ∼1 (Prediction P2, **Figure 2**), consistent with empirical observations ^21^, which is hard to explain otherwise.

While most countries converged to *R*_*e*_ ≈1, they did so at very different levels of average mortality (**Figure 3**). Between-country differences in IFR due to age structure are somewhat predictive of mortality levels (**Figure 4**A and **Table 1**) ^2,5^ while hospital capacity and initial *R*_*0*_ offer limited explanation. Policy response stringency and the exact measures involved have varied across countries and over time^13^, but over the long-term those responses and the corresponding extent of reduction in infectious contacts are not predictive of death rates either (Prediction P3, **Figure 4**D/E/F and **Table 1**). Instead, *responsiveness* becomes the primary explanation for heterogeneity in cases and deaths. Our estimates show that responsiveness varies substantially between different countries, with a strong negative relationship with death rates (Prediction P4, **Figure 5**).

There is no *a priori* conceptual reason these empirical patterns should emerge. Absent endogenous response, *R*_*e*_ would persist well above 1 until the susceptible population is depleted. With exogenous responses, countries could maintain strict NPIs until community transmission is completely suppressed, driving *R*_*e*_ to ∼0 – as a few countries, e.g. China, have done ^29^. In most countries neither scenario has occurred. The convergence of *R*_*e*_ to ∼1 across countries is a key empirical regularity, and the risk response mechanism is sufficient to explain it.

Similarly, intuitive candidate factors such as hospital capacity, IFR, and even policy responses are inadequate to explain wide variation in per capita death rates, despite much evidence of the effect of NPIs on transmission rates ^17–20^. Our model explains this puzzle, showing that *over the long-term*, the driver of variation in mortality is not specific *responses* to perceived risk but rather *responsiveness* to it. With endogenous responses to risk, all communities will sooner or later adopt the responses needed to keep *R*_*e*_ close to 1. Thus, long-term mortality burdens in the face of outbreaks depend less on the specific measures people or countries enact in response, but how severe a situation they will tolerate before they enact them ^32^. Highly responsive countries and communities, which act to reduce transmission when faced with even minor perceived outbreaks, will suffer far fewer deaths. Less responsive countries and communities will wait for those outbreaks to grow larger until they are compelled to act. Thus, they will suffer far more. A corollary implication is that cross-country comparisons may find little association between policy responses and COVID-19 outcomes – but that apparent lack of association does not undermine the necessity of those policy responses.

We have intentionally used an abstract concept of responsiveness to represent the coupling between perceived threat of outbreaks and potential response measures. In reality, responsiveness encompasses several aspects, from public health surveillance capacity to governmental leadership in reacting to outbreak risks to societal willingness to comply with voluntary or mandatory NPIs. Future work could explore psychological, historical, cultural, and institutional factors explaining variations in responsiveness. It is plausible that misunderstanding these dynamics has contributed to suboptimal responses around the world: many communities and policy-makers facing a short-run tradeoff between saving lives and protecting livelihoods have failed to recognize that exponentially growing outbreaks will eventually compel them to act regardless. In the face of future epidemics, a better understanding of these dynamics could help countries toward greater responsiveness, saving many more lives – with little more disruption than they will have to endure anyway.

### Limitations

Our model was designed to highlight how endogenous behavioral responses to risk shape the trajectory and scale of the COVID-19 epidemic across nations. As such, it was intentionally simplified, excluding important features of the pandemic and constraining its ability to replicate observed trajectories. Extensions could incorporate such features as testing and under-reporting, the impact of healthcare capacity on fatality rates, vaccinations, demographics, and SARS-CoV-2 variant emergence, among others.

Another avenue for improvement is the refinement of the endogenous risk response function. First, we did not explore boundary conditions for the risk-response mechanism, e.g. when responsiveness (or IFR) is low enough that even unmitigated outbreaks do not elicit a response. Second, our model does not differentiate between various transmission-reducing policies (e.g. contact tracing, mask mandates, school & business closures, etc.), which can differ both in effectiveness and in how much they disrupt social life or economic activity ^41^. Greater detail would be valuable for understanding how to increase responsiveness, and hence minimize mortality, with minimal disruption. Finally, understanding the drivers of responsiveness differences across communities is a fruitful research topic.

## Data Availability

All models data and analysis code are available at the link below

https://github.com/tseyanglim/CovidRiskResponse.

## Data Sharing

No data were collected specifically for this study; all data are from publicly available databases. All data-processing and analysis code, as well as the full model, its associated files, and results files, are available online at https://github.com/tseyanglim/CovidRiskResponse.

## Author Contributions

TYL curated data, conducted formal analysis, and visualized results. HR conceptualized the study and validated data and analysis. Both TYL and HR developed the model and methodology, conducted investigations, and wrote and edited the manuscript.

## Declaration of Interests

The authors declare no competing interests. No funding was used for this study.

## Online Appendix to Accompany

### S1) Model specification

Our model is an extension of a classic compartmental SEIR model ^1^, which incorporates behavioural responses that endogenously change contact rates as a continuous function of perceived risk.

Populations are divided into four groups: susceptible to the disease (S), exposed but not yet infectious (E), infectious (I), and removed from circulation due to recovery or death (R):

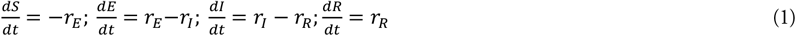

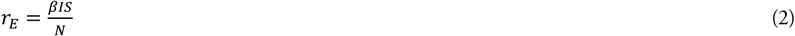

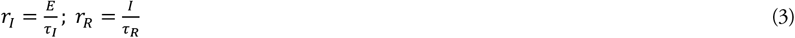

The three main parameters are the daily infectious contacts per case (*β*), the average latent period (*τ*_*I*_), and the average infectious duration (*τ*_*R*_). Susceptible fraction 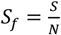 where total population *N* = *S* + *E* + *I* + *R*. The basic reproduction number (*R*_*0*_) is the expected secondary cases from an index case in a fully susceptible population:

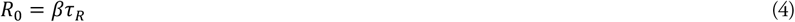

Because outbreaks in first-affected communities change behaviours in others, we call the empirically estimated values for R_0_ *initial reproduction numbers*. Effective reproduction number (*R*_*e*_) then accounts for reductions in secondary cases due to changing susceptible fraction:

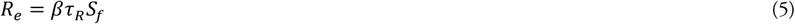

With a constant *β* and an *R*_*0*_ well above one ^2,3^, the basic SEIR model predicts a rapid COVID-19 outbreak that infects most of a population in a few months, reaching herd immunity when a large fraction of the population has been infected ^4^. However, behavioural and policy responses to risk change infectious contact rates (*β*) and may curtail the epidemic well before herd immunity. We thus allow *β* to go below *β*_0_ in response to perceived risk of death (*D*):

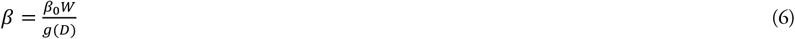

The response function *g* reflects the strength of responses affecting contact rates. *g(D)*=1 indicates pre-pandemic behaviour, whereas full societal lockdown may push *g(D)* to large values, bringing *β* to ∼0. *W* is the seasonal effect of weather on COVID-19 transmission, estimated elsewhere ^5,6^. *g* should be increasing in *D*, but its exact functional form is not critical, so for simplicity we choose a linear form:

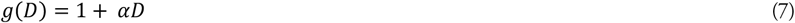

α represents the *responsiveness to risk* of a community. α=0 recovers the basic SIR model, and higher α values indicate a community more sensitive to the perceived risk of death from the disease. *D* is modelled as an exponential average of reported per-capita daily death rate (*d*_*N*_; Equation 8) resulting from an infection fatality rate of *f*, with a time constant of *τ* reflecting the time it takes to perceive and respond to changing risks:

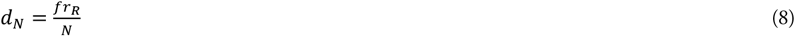

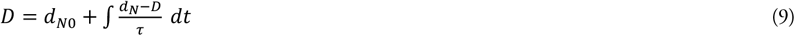

Recognizing that responding to growing risks and relaxing responses based on declining risks may occur on different timescales, we allow for an asymmetric adjustment time *τ* for increasing vs. decreasing perceived risk:

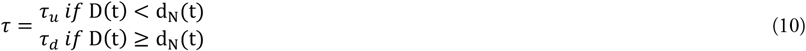

To capture potential changes in risk-response relationship over time (e.g. due to adherence fatigue), we allow responsiveness to risk (α) to vary linearly with time:

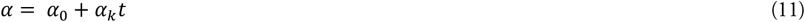

This model includes 8 parameters (*τ*_*I*_, *τ*_*R*_, *β*_0_, *α*_0_, *α*_*k*_, *f, τ*_*u*_, *τ*_*d*_) as well as one time-varying input *W*. Based on prior literature, we specify *τ*_*I*_ = 4 and *τ*_*R*_=10 days ^7,8^. We calculate base IFR for each country based on age distribution ^9^ (see S3.a)), and use prior estimates for *W* ^5,6^. The remaining five parameters (*β*_0_, *α*_0_, *α*_*k*_, *τ*_*u*_, *τ*_*d*_) are estimated. In addition, for each country, we estimate the arrival time of the first patient (*t*_*Z*_), yielding a parameter vector ***θ*** with 6 unknown parameters per country.

#### S1.a) Accounting for under-ascertainment

Under-ascertainment is a substantial challenge for models of COVID-19 transmission. To account for under-ascertainment of cases, our model distinguishes between reported daily infections and true underlying infection rates (*r*_*E*_) for each country:

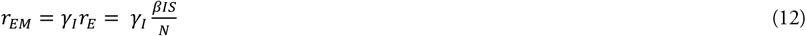

The subscript *M* denotes reported (rather than true) values, and *γ*_*I*_ is the ratio of reported to true infections. To keep the model simple, we side-step needing to estimate ascertainment by approximating reported infection rates *r*_*EM*_ based on recent reported infections *I*_*M*_, assuming that current infections are under-reported to the same degree as infections over the last few weeks:

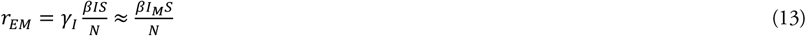

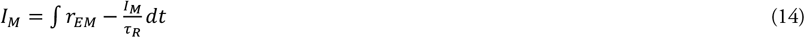

This approximation holds as long as *γ*_*I*_ is relatively stable on the timescale of the disease duration (∼two weeks), even if it changes over longer time horizons.

### S2) Estimation method

Our model yields an expected number of new reported infections (*r*_*EM*_) and per-capita death rate (*d*_*N*_) for each day for each country. Those expected numbers are specified in Equations 13 and 8 in S1), as a function of several known and unknown parameters and the state of the model prior to the current date. We simulate the system of differential equations captured in the model to calculate those outcomes over time. We estimate the vector ***θ*** of unknown parameters for each country by maximum likelihood, identifying the vector 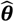 that maximises the likelihood of observing the true reported infections and deaths *y*_*vt*_ (where *t* is time (day) and *v* ∈ [*i, d*] denotes the series of infections or deaths) given 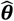.

We use a negative binomial [log-]likelihood function for both cases and deaths:

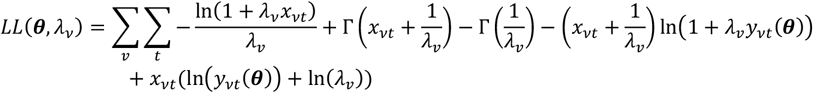

Where *x*_*vt*_ is predicted infections or deaths (analogous to *y*_*vt*_), *λ*_*v*_ are the scaling factors for the likelihood function, and Γ(z) represents the natural logarithm of the generalized factorial function for *z* − 1 (i.e. Γ(*z*+ 1) *z*! for integer *z*).

The vector ***θ*** includes 6 unknown parameters, as listed in **Table S1**. We also estimate the negative binomial scaling factors (*λ*_*i*_, *λ*_*d*_), leading to a total of 8 estimated parameters for each country. The NRR version of the model fixes the two responsiveness parameters (*α*_0_ and *α*_*k*_) at 0, and thus has 6 estimated parameters per country.

**Table S1.** Estimated parameters with allowed ranges.

| Parameter | Symbol | Range | Units |
| --- | --- | --- | --- |
| Initial infectious contact rate | $\beta_0$ | 0.1-4.0 | People/person/day |
| Initial responsiveness | $\alpha_0$ | 0.001-1000 | Dimensionless |
| Responsiveness scaling factor | $\alpha_k$ | 1.1E-03-1.1* | Dimensionless |
| Time to increase perceived risk | $\tau_u$ | 1-100 | Days |
| Time to reduce perceived risk | $\tau_d$ | 10-400 | Days |
| Patient Zero arrival time** | $t_z$ | 1-140 | Day |
| Likelihood scaling factor (infections) | $\lambda_i$ | 1E-06-1 | Dimensionless |
| Likelihood scaling factor (deaths) | $\lambda_d$ | 1E-12-0.6*** | Dimensionless |

\* For ease of computation, *Responsiveness scaling factor* is defined relative to $\alpha_0$ as $\pm 0.5 \times \alpha_0 / t_{max}$ (=455)
\*\* *Patient Zero arrival time* is expressed in days from the start of 2020
\*\*\* To avoid a potential computational problem in which the estimation algorithm ignores deaths relative to infections in the likelihood function, the scaling factor for deaths $\lambda_d$ is defined relative to $\lambda_i$ as $1E-06-0.6 \times \lambda_i$

We estimate the model separately for each country. Separating countries significantly speeds up estimation and makes it feasible to conduct the full analysis with limited computing resources. For each country, we estimate the parameter vector ***θ*** using the Powell direction search method implemented in Vensim™ DSS simulation software, restarting the optimization at 20 random points in the feasible parameter space. Overall, estimation for all 131 countries takes approximately 12 hours when compiled and parallelized on a 48-core Windows 10 server.

### S3) Data sources & preparation

Data on daily confirmed cases and deaths come from the OurWorldInData (OWID) global COVID-19 database ^10^, which draws on the Johns Hopkins University CSSE COVID dashboard ^11^. The CSSE dashboard in turn aggregates its data primarily from official sources such as the US Centres for Disease Control and Prevention (CDC), the European CDC, the World Health Organization, and national health ministries, updating at least daily.

We use OWID’s 7-day rolling averages for new cases (‘new_cases_smoothed’) and deaths per million population (‘new_deaths_smoothed_per_million’). COVID-19 case and death reporting data show strong weekly cycles in many countries, as well as occasional anomalous spikes due to e.g. irregularities in test reporting or redefinitions by government statistical agencies; using the rolling average data smooths out these cycles, which we are not attempting to model here, to better reflect underlying trends.

Our analysis includes all countries in the dataset with at least 10000 cumulative cases reported, and at least 20 days of data, by 31 Mar 2021. We exclude countries with fewer than 10000 cumulative cases to avoid skewing the results with outliers and ensure robust estimation. In total, 131 countries meet these criteria by 31 Mar 2021. As discussed in the main text, we restrict the estimation period to 31 Dec 2019 to 31 Mar 2021 to avoid the confounding impacts of vaccination and new variants, which are beyond the scope of this model.

For countries included, we utilise data starting from the date when they exceed 100 cumulative cases reported. Excluding early data entails a trade-off. Excluding it makes estimating the true basic reproduction number (*R*_*0*_) more difficult –after forceful outbreaks in the first countries, most others adopted various responses that brought *R*_*e*_ down below its pre-pandemic level (*R*_*0*_). Furthermore, excluding the early data may cut out the initial dynamics of infection. As a result, our estimated values for initial reproduction number are likely underestimates of basic reproduction number, and thus the *g* estimates may tend to be lower than the true changes in the contact rates compared to pre-pandemic levels. On the other hand, many of the early cases reported in most countries were due to travellers, and often identified and isolated early on. The data during this earliest ‘importation’ stage therefore may not accurately reflect the community transmission dynamics we are modelling. Rapid changes in testing coverage also impact our ability to assume ascertainment rates are stable in the *τ* time horizon as needed in our derivations (see Equation 13). We selected the 100 case cut-off to balance reasonably estimating *R*_*0*_ against correctly reflecting transmission dynamics rather than travel networks, which are out of scope for this model.

For analysis of results (but not model estimation), we use several other country statistics such as GDP, hospital beds per thousand people, median age, and so on, with data as compiled by OWID^1 10^. We also use government response stringency data ^12^ and independent estimates of *R*_*e*_ ^13^ compiled by OWID and available through their COVID-19 data hub. The full list of additional variables used and corresponding OWID data codes is in **Table S2** below.

For mobility data we use Google’s COVID-19 Community Mobility Reports^2^, accessed via OWID^3^ to provide consistent mapping for country names to other data we use. These data reflect changes in numbers of visitors relative to pre-pandemic levels, controlling for weekly (but not longer seasonal) cycles. We use the averaged value of percentage changes in visits to two categories of locations – workplaces, and retail & recreation venues – which best reflect normal everyday economic activity.

Data are downloaded and processed with Python 3 code, using Pandas and NumPy packages. For the full data processing code, see https://github.com/tseyanglim/CovidRiskResponse.

**Table S2.**
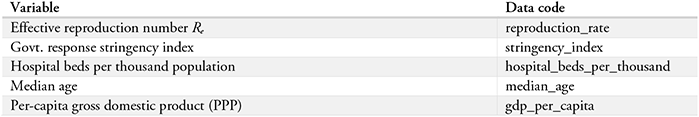
Additional variables drawn from OWID dataset and variable codes.

#### S3.a) IFR calculation

Age strongly influences IFR, with older patients far more likely to die of COVID-19 ^9^. To account for the impact of demographic differences on fatality rates and improve model estimation, we therefore calculate country-specific IFRs based on each country’s age structure.

We use data from the World Bank’s World Development Indicators ^14^ on the age distribution of each country’s population in 10-year age strata to calculate an age-weighted average of the IFRs for COVID patients by 10-year age group estimated in ^9^. The resulting demography-adjusted country-specific IFRs range from 0.14% (Uganda) to 1.51% (Japan), with a mean of 0.54% and median of 0.44% (Lebanon). For the handful of countries for which up-to-date demographic data are unavailable, we use a baseline IFR of 0.50%.

### S4) Full results

#### S4.a) Model fit to data

**Table S3** shows goodness-of-fit statistics for simulated infection and death rates against data (median across all 131 countries) for the base model and the no-risk-response (NRR) version, while **Figure S1** and **Figure S2** show individual country fits to data for simulated infection rates (**Figure S1**) and death rates (**Figure S2**) for all 131 countries. Solid lines show model-generated daily infection or death rates, while dotted lines show 7-day rolling average infection rates from OWID.

For infections, the correspondence between model and data is very close for most countries, with a few outliers bringing down the quality of fit a bit; over the full sample of 131 countries, however, R^2^ for infections against data is still 0.86, while the mean absolute errors normalized by mean (MAEN) are 25%. The quality of fit should not come as a total surprise: by calculating “reported” infections based on accumulated reported infection rates (see Equation 13), the model is in effect starting the prediction of cases from a data-informed anchor, rather than a purely simulated value. This formulation reduces error due to the potentially accumulated gap between simulation and data, thus improving fit ^15^. Nevertheless, the risk response function still substantially improves quality of fit (**Table S3**). In contrast, for death rates we simply use the simulated ‘actual’ cases, which are far more inaccurate, and thus the quality of fit for reported deaths is worse than for infections (**Figure S2**), with an R^2^ of 0.27 and MAEN of 100%. Limited quality of fit is unsurprising given the simplicity of the model and the fact that we calculated deaths based on simulated trajectories, rather than data, and death rates vary by 2-3 orders of magnitude across countries; more importantly, as with infections, the risk response function improves fit substantially, reducing MAEN by an order of magnitude (**Table S3**).

**Table S3.** Goodness-of-fit statistics for infection and death rates, for model with / without endogenous behavioural risk response.

|  | Infections |  | Deaths |  | Weighted Avg. |  |
| --- | --- | --- | --- | --- | --- | --- |
| | $R^2$ | MAEN | $R^2$ | MAEN | $R^2$ | MAEN |
| With risk response | 0.86 | 25% | 0.27 | 100% | 0.56 | 66% |
| No risk response | 0.55 | 55% | 0.15 | 1042% | 0.36 | 551% |

**Figure S1.**
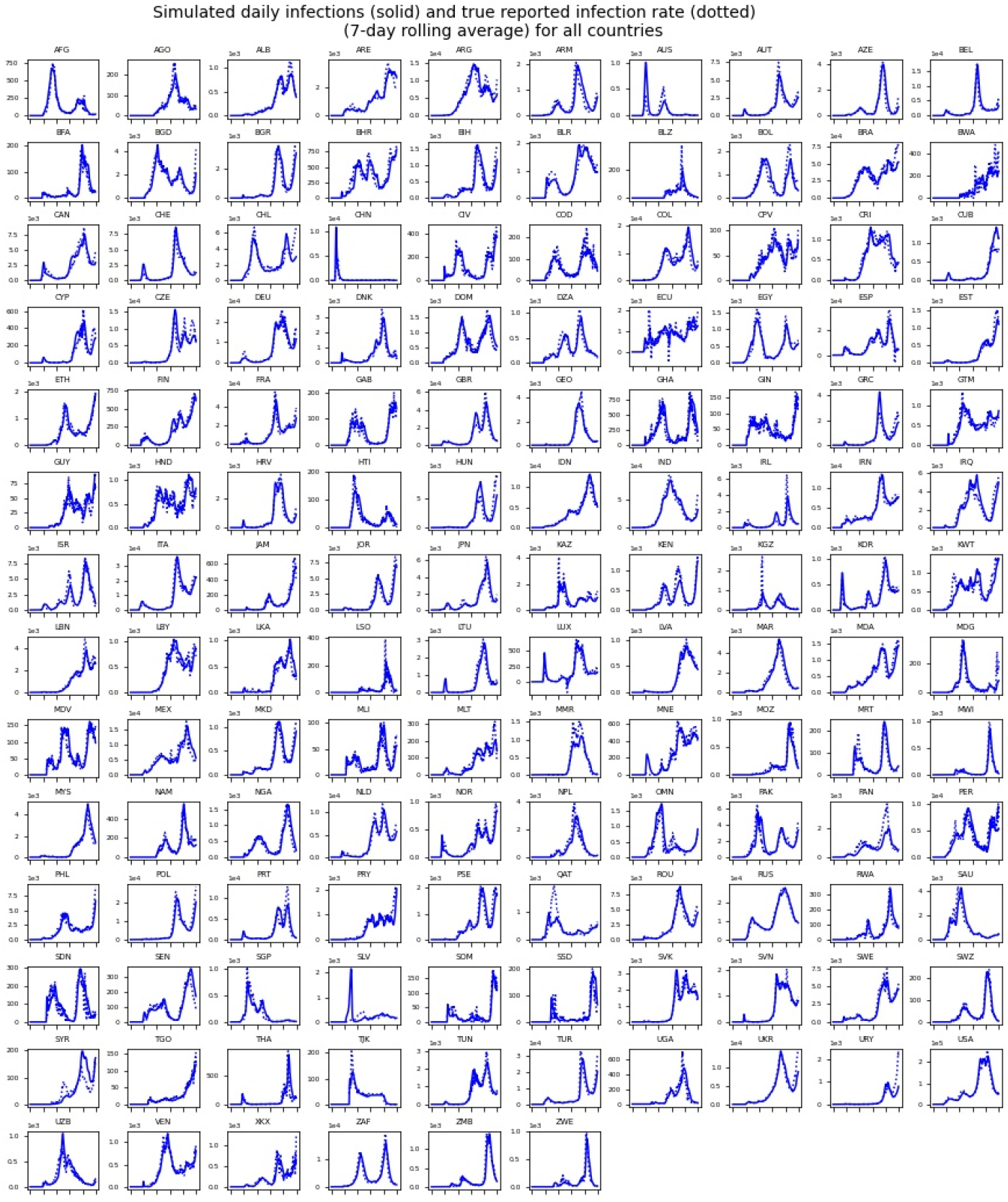
Model output fit to data for reported daily infection rates for all countries.

**Figure S2.**
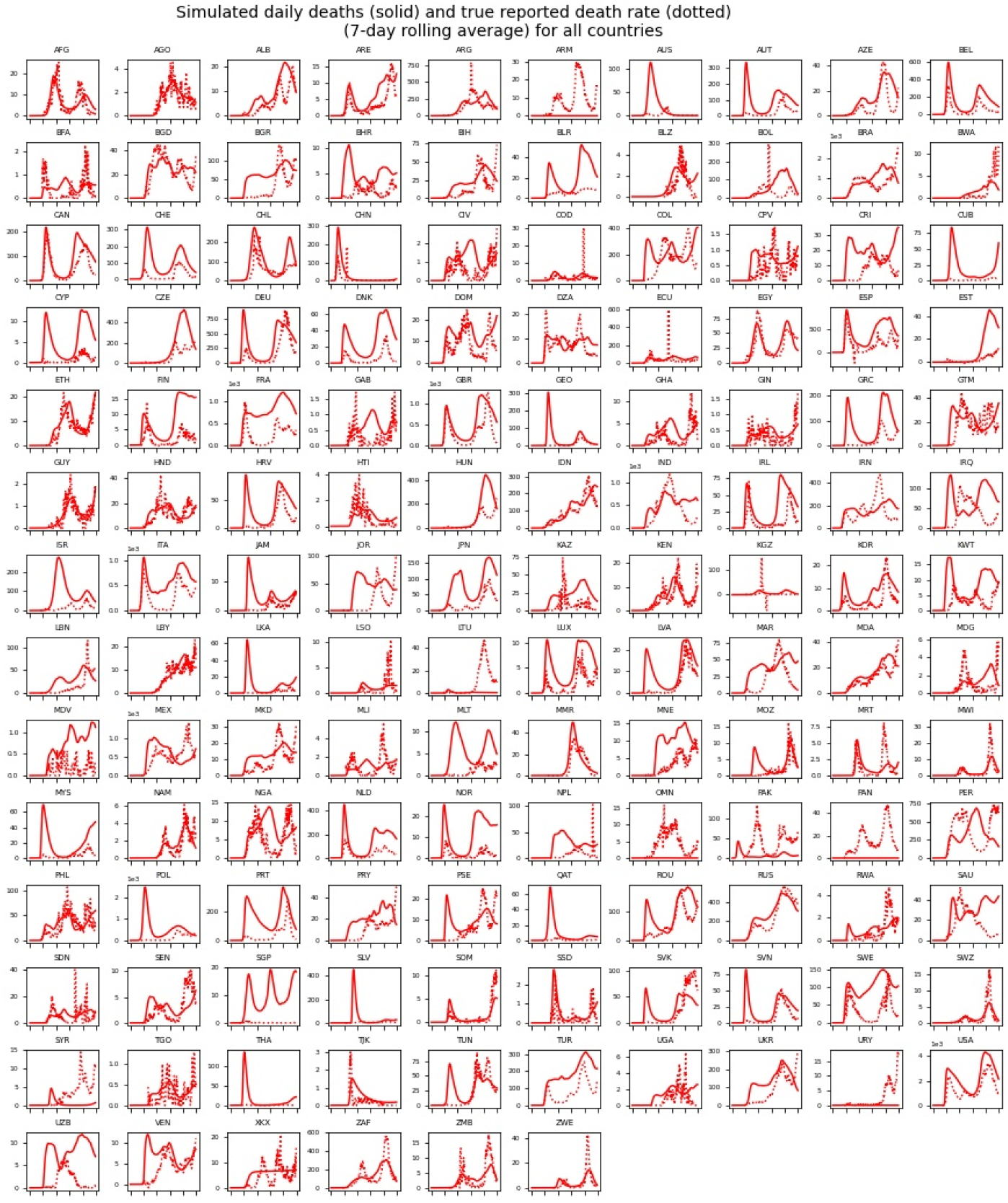
Model output fit to data for reported daily death rates per million people for all countries.

#### S4.b) Correlations with additional potential explanatory variables

**Figure S3** shows additional correlations (or lack thereof) between reported per-capita death rates (averaged over six months up to 31 Mar 2021) and various country characteristics. Across countries, death rates are actually *positively* correlated with per-capita GDP, hospital capacity, and median age (**Figure S3**A-C), indicating that these factors likely do not drive mortality variation across countries. Intuitively, wealth / GDP and hospital capacity should be *negatively* correlated with death rates, and this pattern is seen within some countries and on shorter timescales ^16^. The counterintuitive correlation is likely driven primarily by median age, as wealthier countries with greater healthcare capacity tend to have longer lifespans and thus older populations more vulnerable to COVID-19 mortality. Indeed, when death rates are adjusted for age (normalising by their respective age-adjusted IFRs relative to average IFR across countries), these correlations are greatly reduced or disappear entirely (**Figure S3**D-F).

**Figure S3.**
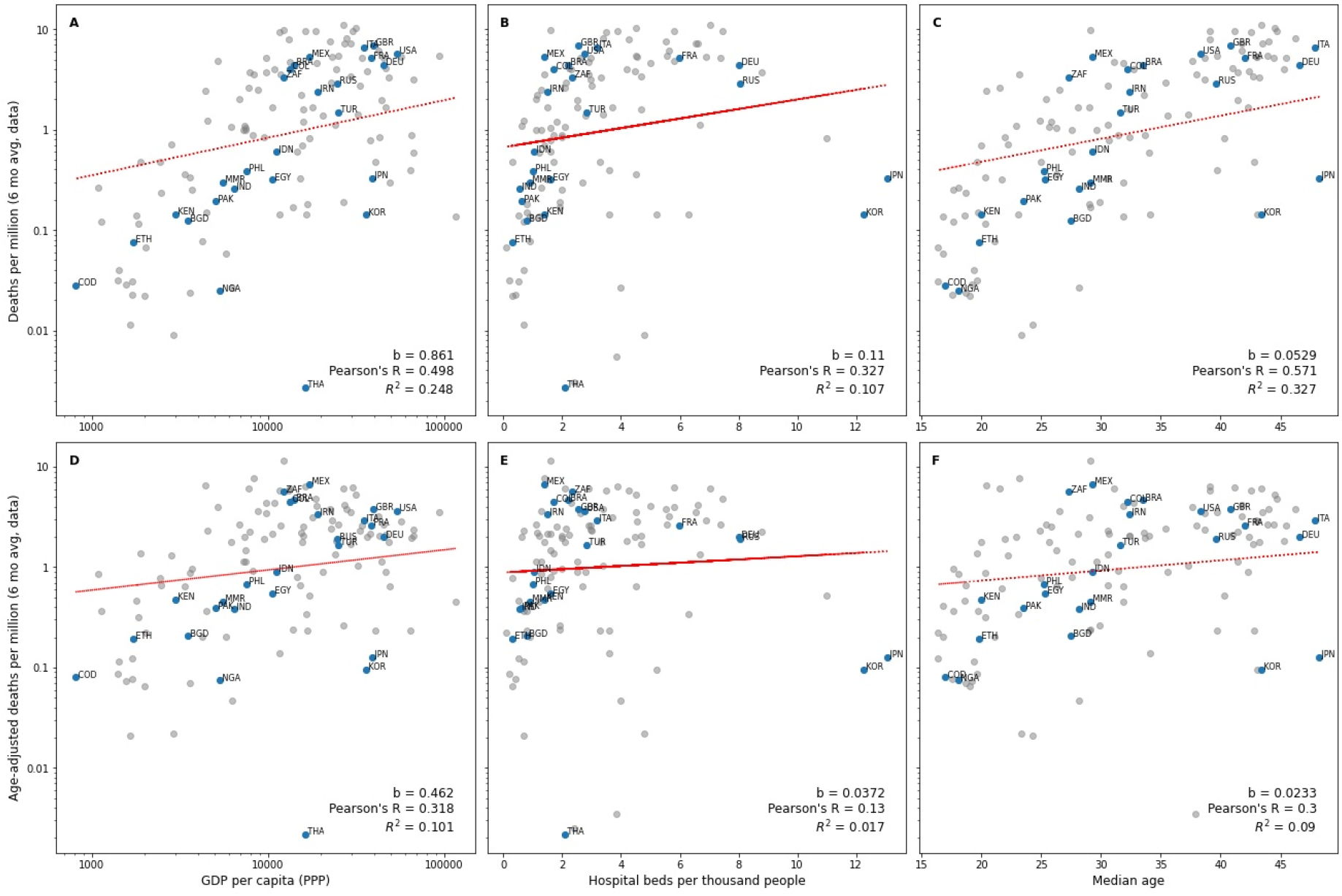
Reported death rates per million people correlated with various factors. A) GDP per capita (PPP); B) hospital beds per thousand people; C) median age. D)-F) show correlations of age-adjusted death rates per million (normalized by age-adjusted IFR) with the same factors as A)-C) respectively.

**Figure S4** shows estimated death rates per million against change in visits to retail & recreation venues and workplaces respectively, relative to pre-pandemic levels. The two categories separately show similar negative correlation to the combined mobility index, as shown in the main text.

**Figure S4.**
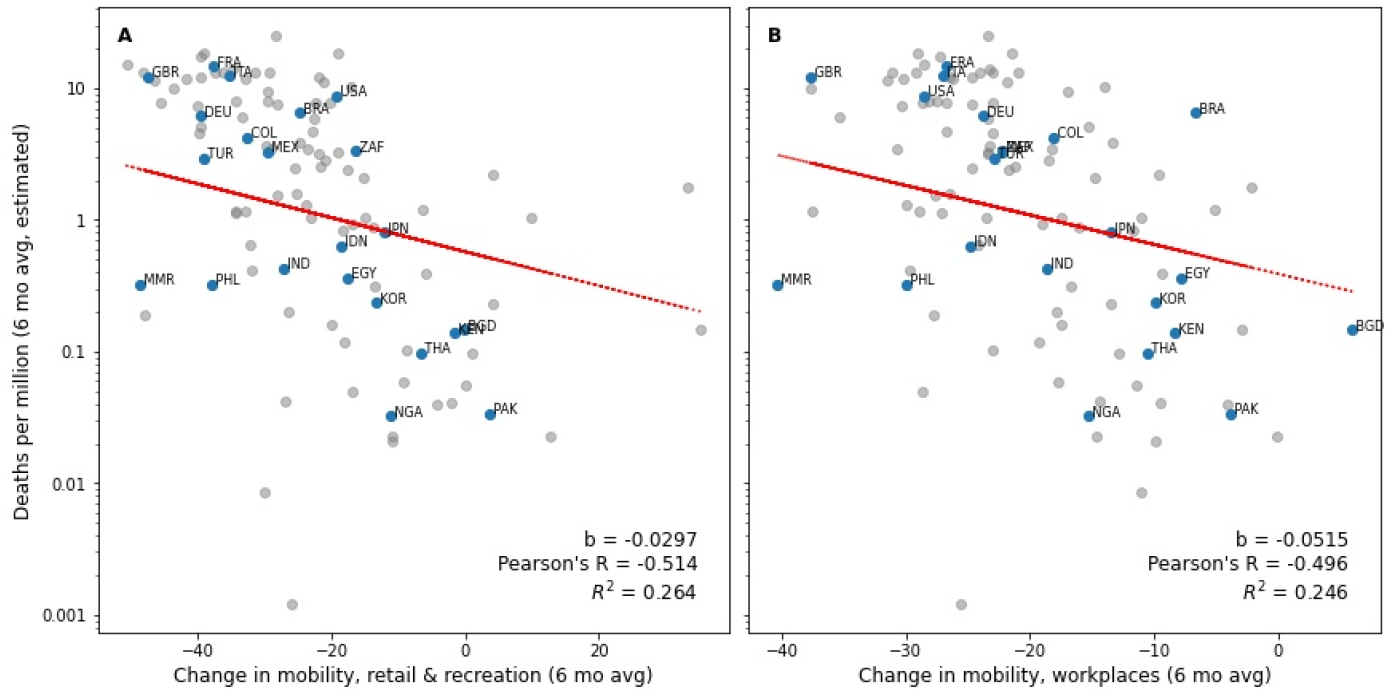
Estimated death rates per million correlated with change in mobility relative to pre-pandemic levels for A) retail & recreation venues and B) workplaces.

#### S4.c) Parameter estimates

**Table S4** shows summary statistics across all countries for estimated model parameters. For the full table of parameter estimates by country, see https://github.com/tseyanglim/CovidRiskResponse.

**Table S4.** Summary statistics of estimated parameter values.

| Parameter | Symbol | Mean | Median | StDev |
| --- | --- | --- | --- | --- |
| Initial infectious contact rate | $\beta_0$ | 1.81 | 1.49 | 1.35 |
| Initial responsiveness | $\alpha_0$ | 82.7 | 5.28 | 200 |
| Responsiveness scaling factor | $\alpha_k$ | -0.0234 | -0.00136 | 0.183 |
| Time to increase perceived risk | $\tau_u$ | 18 | 3.38 | 28.1 |
| Time to reduce perceived risk | $\tau_d$ | 103 | 28.5 | 136 |
| Patient Zero arrival time* | $t_z$ | 65.5 | 63 | 31.9 |
| Likelihood scaling factor (infections) | $\lambda_i$ | 0.478 | 0.451 | 0.334 |
| Likelihood scaling factor (deaths) | $\lambda_d$ | 0.366 | 0.32 | 0.279 |

\* Patient Zero arrival time is expressed in days from the start of 2020

#### S4.d) Predictor variable correlations & variance inflation factors

**Table S5** shows correlations between all predictor variables used in the full death rate regression (**Table 1** of main manuscript), as well as variance inflation factors (VIFs); all VIF values are < 3, indicating acceptably low multicollinearity between predictors.

**Table S5.** Correlation matrix and variance inflation factors for predictor variables in full regression.

|  | log_avg_Sensitivity<br>Alpha_180 | log_IFR | log_gdp_per_capita | hosp_beds | R0_est | hist_str_180 | Variance<br>Inflation<br>Factor |
| --- | --- | --- | --- | --- | --- | --- | --- |
| log_avg_Sensitivity Alpha_180 | 1 | -0.567 | -0.556 | -0.369 | -0.141 | -0.336 | 1.73 |
| log_IFR | -0.567 | 1 | 0.654 | 0.630 | 0.416 | 0.297 | 2.66 |
| log_gdp_per_capita | -0.556 | 0.654 | 1 | 0.447 | 0.391 | 0.376 | 2.10 |
| hosp_beds | -0.369 | 0.630 | 0.447 | 1 | 0.357 | 0.019 | 1.82 |
| R0_est | -0.141 | 0.416 | 0.391 | 0.357 | 1 | 0.213 | 1.34 |
| hist_str_180 | -0.336 | 0.297 | 0.376 | 0.019 | 0.213 | 1 | 1.30 |

### S5) Model equations listing

1. AdjIFR[Rgn] = GET VDF CONSTANTS(‘InputConstants.vdf’, ‘AdjIFR[Rgn]’, 1)
2. alp[Rgn,Infection] = 0.01
3. alp[Rgn,Death] = alp[Rgn,Infection] * alpratio[Rgn]
4. Alpha 0[Rgn] = 1
5. Alpha F[Rgn] = Alpha 0[Rgn] * Alpha Rel[Rgn]
6. Alpha Rel[Rgn] = 1
7. alpratio[Rgn] = 0.5
8. Asymptomatic fraction = 0.5
9. BaseIFR = 0.005
10. Beta[Rgn] = 0.45
11. Constant Data File:IS:’StatePopulations.vdf’
12. CRW[Rgn]:INTERPOLATE:
13. DataFlow[Rgn,Infection]:RAW:: = if then else(new cases[Rgn] = :NA:, :NA:, new cases[Rgn])
14. DataFlow[Rgn,Death]:RAW:: = if then else(new cases[Rgn] = :NA:, :NA:, new deaths[Rgn])
15. DataFlowExport[Rgn,Series] = if then else(Time < DataStartTime[Rgn,Series], -999, DataFlowInterpolated[Rgn,Series])
16. DataFlowInterpolated[Rgn,Infection]:INTERPOLATE:: = new cases[Rgn]
17. DataFlowInterpolated[Rgn,Death] = new deaths[Rgn]
18. DataStartTime[Rgn,Infection] = INITIAL(GET DATA FIRST TIME(new cases[Rgn]))
19. DataStartTime[Rgn,Death] = INITIAL(GET DATA FIRST TIME(new deaths[Rgn]))
20. DataToInclude[Series] = 1,1
21. Dead[Rgn] = INTEG (Deaths[Rgn],0)
22. DeathDataActive = if then else(Time>Last Estimation Time,0,1)*UseDeathData
23. Deaths[Rgn] = Removals[Rgn]*IFR[Rgn]
24. DeathStartValue[Rgn] = GET DATA AT TIME(new deaths[Rgn], :NA:)
25. Di[Rgn,Series] = DataFlow[Rgn,Series]
26. DiscountRate = 0
27. DiseaseDuration = 10
28. E[Rgn] = INTEG (Exposure[Rgn]+Patient Zero Arrival[Rgn]-Onset[Rgn],0)
29. Early Death Penalty[Rgn] = if then else (Time < DataStartTime[Rgn,Death], (1-TimeToIncludeFromData[Rgn,Death])*Mu[Rgn,Death] -DeathStartValue[Rgn], 0)
30. eps = 0.001
31. Expected Reported Infections[Rgn] = IMeas[Rgn]*Indicated Attack Rate[Rgn]*Susceptible Fraction[Rgn]
32. Exposure[Rgn] = I[Rgn]*Indicated Attack Rate[Rgn]*Susceptible Fraction[Rgn]
33. FINAL TIME = 455
34. I[Rgn] = INTEG (Onset[Rgn]-Deaths[Rgn]-Recovery[Rgn],0)
35. IFR[Rgn] = INITIAL(if then else(AdjIFR[Rgn] = -1, BaseIFR, AdjIFR[Rgn]))
36. IMeas[Rgn] = INTEG (onsetMeas[Rgn]-RemovMeas[Rgn],0)
37. Impact of perceived risk on attack rate[Rgn] = 1/(1+(Perceived death rate[Rgn]*Sensitivity Alpha[Rgn])^PWRisk[Rgn])
38. InclusionThreshold[Rgn,Series] = INITIAL(Population[Rgn]*Threshold[Series])
39. Indicated Attack Rate[Rgn] = Beta[Rgn]*Impact of perceived risk on attack rate[Rgn]*Weather Effect on Transmission[Rgn]
40. Indicated Death Rate[Rgn] = if then else(DeathDataActive = 0,Deaths[Rgn], DataFlow[Rgn,Death]) /Population[Rgn]*Normalized population size
41. Infection reporting fraction[Rgn] = ZIDZ(onsetMeas[Rgn], Onset[Rgn])
42. INITIAL TIME = 1
43. Last Estimation Time = 1000
44. LastStart[Series] = 200,200
45. Limit Prior Time = 0
46. Mu[Rgn,Series] = Max (eps, Outputs[Rgn,Series])
47. NBL1[Rgn,Series] = if then else (DataFlow[Rgn,Series] = 0, -ln (1+ alp[Rgn,Series] * Mu[Rgn,Series]) / alp[Rgn,Series], 0)
48. NBL2[Rgn,Series] = if then else (DataFlow[Rgn,Series] > 0, GAMMA LN (Di[Rgn,Series] + 1 / alp[Rgn,Series]) -GAMMA LN (1 / alp[Rgn,Series]) - GAMMA LN (Di[Rgn,Series] + 1) - (Di[Rgn,Series] + 1 / alp[Rgn,Series]) * ln (1 + alp[Rgn,Series] * Mu[Rgn,Series]) + Di[Rgn,Series] * (ln (alp[Rgn,Series]) + ln (Mu[Rgn,Series])), 0)
49. NBLLFlow[Rgn,Series] = (NBL1[Rgn,Series] + NBL2[Rgn,Series]#x002A;DataToInclude[Series]*TimeToInclude[Rgn,Series]*(Time/FINAL TIME)^DiscountRate
50. new cases[Rgn]:RAW:
51. new deaths[Rgn]:RAW:
52. Normalized population size = 1e+06
53. Onset[Rgn] = E[Rgn]/Time to onset Te[Rgn]
54. onsetMeas[Rgn] = if then else(Time < DataStartTime[Rgn,Infection], 0, DataFlowInterpolated[Rgn,Infection])
55. Outputs[Rgn,Infection] = Expected Reported Infections[Rgn]
56. Outputs[Rgn,Death] = Deaths[Rgn]
57. Patient zero = 1
58. Patient Zero Arrival[Rgn] = if then else (Time < Patient Zero Arrival Time[Rgn] :AND: Time + TIME STEP > = Patient Zero Arrival Time[Rgn], Patient zero / TIME STEP, 0)
59. Patient Zero Arrival Time[Rgn] = 100
60. Perceived death rate[Rgn] = INTEG ((Indicated Death Rate[Rgn]-Perceived death rate[Rgn])/Time to Perceive Risk[Rgn],Indicated Death Rate[Rgn])
61. Population[Rgn] = GET VDF CONSTANTS(‘InputConstants.vdf’, ‘Population[Rgn]’, 1)
62. PWRisk[Rgn] = 1
63. Re[Rgn] = Indicated Attack Rate[Rgn] * Susceptible Fraction[Rgn] * Time to removal Tr[Rgn]
64. Recovered[Rgn] = INTEG (Recovery[Rgn],0)
65. Recovery[Rgn] = Removals[Rgn]*(1-IFR[Rgn])
66. Removals[Rgn] = I[Rgn]/Time to removal Tr[Rgn]
67. RemovMeas[Rgn] = IMeas[Rgn]/Time to removal Tr[Rgn]
68. Reporting fraction penalty[Rgn] = Max(IMeas[Rgn] - I[Rgn] * Asymptomatic fraction, 0) / 10
69. S[Rgn] = INTEG (-Exposure[Rgn],Population[Rgn] -1)
70. SAVEPER = 1
71. Sensitivity Alpha[Rgn] = Max(Alpha 0[Rgn]+Time/FINAL TIME * (Alpha F[Rgn]-Alpha 0[Rgn]), 0)
72. Sensitivity to Weather = 2.64
73. Series:Infection,Death
74. Shft:(S1-S20)
75. Susceptible Fraction[Rgn] = S[Rgn]/Population[Rgn]
76. Threshold[Series] = 1e-06,5e-08
77. TIME STEP = 0.25
78. Time to increase risk[Rgn] = 60
79. Time to onset Te[Rgn] = 4
80. Time to Perceive Risk[Rgn] = (if then else(Indicated Death Rate[Rgn]>Perceived death rate[Rgn],Time to increase risk[Rgn],Time to reduce risk[Rgn]))
81. Time to reduce risk[Rgn] = 60
82. Time to removal Tr[Rgn] = DiseaseDuration
83. TimeToInclude[Rgn,Series] = if then else(Time< = Last Estimation Time :AND: Time>Last Estimation Time*Limit Prior Time-100,1,0)*TimeToIncludeFromData[Rgn,Series]
84. TimeToIncludeFromData[Rgn,Series] = if then else(DataFlow[Rgn,Series]>InclusionThreshold[Rgn,Series] :OR: Time>LastStart[Series],1,0)
85. UseDeathData = 0
86. Weather Effect on Transmission[Rgn] = if then else(CRW[Rgn] = :NA:, 1, CRW[Rgn]^Sensitivity to Weather)
87. XErrAbs[Rgn] = ABS(Di[Rgn,Death]-Outputs[Rgn,Death])*if then else(Time>Last Estimation Time,1,0)

## Footnotes

1 https://raw.githubusercontent.com/owid/covid-19-data/master/public/data/owid-covid-data.csv

2 https://www.google.com/covid19/mobility

3 https://ourworldindata.org/covid-mobility-trends

## Notes

### Competing Interest Statement

The authors have declared no competing interest.

### Funding Statement

This research used no funding

### Author Declarations

Only public data sources used

### Summary of Updates

Refocus the analysis on explaining large variations in mortality across countries

